# Mechanical Dyssynchrony and Perfusion Heterogeneity Predict Adverse LV Remodeling in Patients with and without LBBB

**DOI:** 10.64898/2026.01.09.26343726

**Authors:** Simone Cristina Soares Brandão, Lee Joseph, Jenifer M. Brown, Diana Lopez, Mark Lemley, Giselle Ramirez, Paul Kavanagh, Serge D. Van Kriekinge, Joanna Liang, Valerie Builoff, Sanjay Divakaran, Brittany Weber, Jon Hainer, Sylvain Carre, Ron Blankstein, Sharmila Dorbala, Viet T. Le, Steve Mason, Stacey Knight, Panithaya Chareonthaitawee, Samuel Wopperer, Thomas L. Rosamond, Daniel S. Berman, Damini Dey, Marcelo F. Di Carli, Piotr J. Slomka, Daniel M. Huck

## Abstract

**Aims:** Left bundle branch block (LBBB) is associated with mechanical dyssynchrony, heterogeneous perfusion, and adverse left ventricular (LV) remodeling. However, not all patients with LBBB develop cardiomyopathy, and dyssynchrony can occur without conduction defects. The role of microvascular dysfunction remains uncertain. We aimed to assess how mechanical dyssynchrony and perfusion heterogeneity relate to LV remodeling and function in patients with and without LBBB.

**Methods and results:** We retrospectively analyzed 233 patients with isolated LBBB and 932 matched controls who underwent PET myocardial perfusion imaging, assessing mechanical dyssynchrony (phase entropy), myocardial blood flow (MBF), coronary vascular resistance (CVR), myocardial flow reserve (MFR), septal-to-lateral MBF ratio (SLR) for perfusion heterogeneity, LV volumes, and ejection fraction (EF). Compared to controls, patients with LBBB had greater dyssynchrony (56% vs. 40%), larger LV volumes, and lower EF (54% vs. 67%) (all *p*<0.001), and had higher stress CVR (37 vs. 34 mmHg/mL·min⁻¹·g⁻¹), lower stress MBF (2.4 vs. 2.6 mL/min/g), reduced MFR (2.4 vs. 2.6), and lower SLR (0.95 vs. 1.00) (all *p*<0.05).

Among patients with dyssynchrony, SLR<1.0 identified those with more adverse remodeling. In multivariable regression, phase entropy and SLR independently predicted LV volumes and EF, with adverse effects of SLR reduction amplified in LBBB (interaction *p*<0.01). In the Cox proportional hazards analysis, phase entropy (HR:1.02, *p*=0.01), MFR (HR:0.62, *p*<0.001), and LVEF (HR:0.97, *p*<0.001) were independently associated with mortality and heart failure hospitalization, whereas LBBB was not.

**Conclusions:** Mechanical dyssynchrony and perfusion heterogeneity independently predict adverse LV remodeling, irrespective of LBBB. Integrated imaging enhances cardiomyopathy stratification.

## INTRODUCTION

Left bundle branch block (LBBB) is a conduction disturbance that causes dyssynchronous electromechanical activation of the left ventricle (LV), leading to mechanical inefficiency and, in some patients, to structural remodeling and progression to cardiomyopathy (1–3). Although LBBB is traditionally recognized by its electrocardiographic features, not all patients with LBBB experience significant mechanical dyssynchrony or adverse LV remodeling, and not all mechanical dyssynchronies lead to LV dysfunction, suggesting that additional factors modulate the natural history of the myocardial response (4). One such potential mechanism is alterations in coronary microvascular perfusion that may be associated with mechanical dyssynchrony and co-existing cardiometabolic risk factors. Prior studies suggested that septal hypoperfusion in LBBB results from autoregulatory reductions in local blood flow secondary to diminished oxygen demand in the early-activated myocardium (5–8). We hypothesize that such alterations in myocardial perfusion may serve as a marker of impaired coupling between electrical activation and contractile performance, thereby contributing to adverse remodeling and myocardial dysfunction.

Accordingly, we sought to evaluate the association between the degree of mechanical dyssynchrony and regional perfusion heterogeneity in patients with and without LBBB, and to assess the interrelationship between myocardial dyssynchrony and impaired myocardial perfusion parameters with changes in LV remodeling and function (**Central Illustration**).

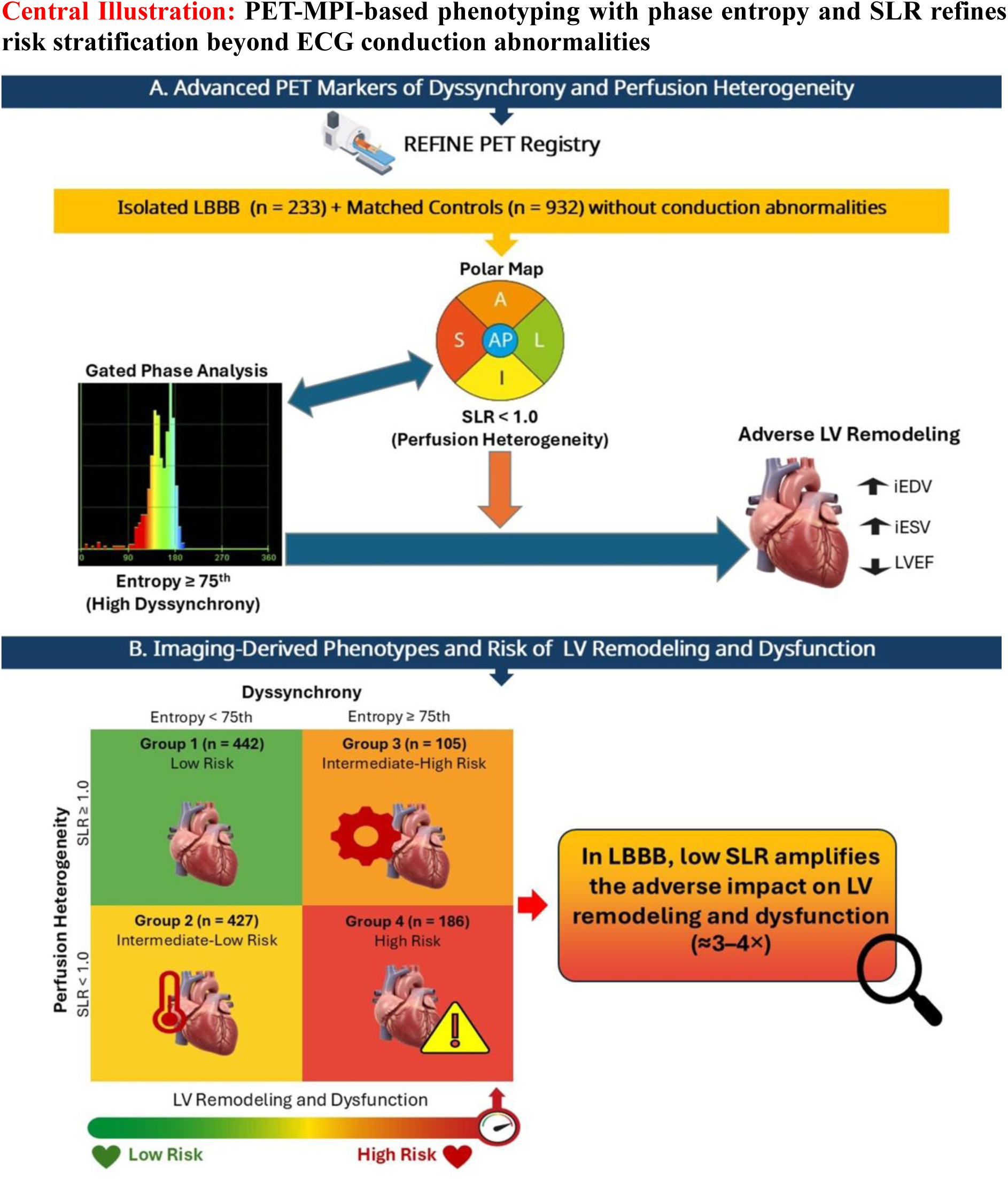
Phase entropy (from gated PET MPI) quantifies the dispersion of LV contraction phases, reflecting mechanical dyssynchrony. High dyssynchrony was defined as phase entropy ≥75th percentile (≈52%) from the overall cohort (LBBB + controls). Combined assessment of phase entropy and SLR identifies dyssynchrony and perfusion heterogeneity, enabling early phenotyping and improved prediction of LV remodeling and dysfunction. iEDV = Indexed End-Diastolic Volume; iESV = Indexed End-Systolic Volume; LBBB = Left Bundle Branch Block; LV = Left Ventricle; LVEF = Left Ventricular Ejection Fraction; MBF = Myocardial Blood Flow; MPI = Myocardial Perfusion Imaging; PET = Positron Emission Tomography; SLR = Septal-to-Lateral (myocardial blood flow) Ratio.

## METHODS

### Study Population

We retrospectively analyzed patients enrolled in the multicenter REFINE PET registry (9). From a total cohort of 17,887 patients, we identified 233 patients with isolated LBBB without perfusion abnormalities (summed stress score <4). LBBB was identified based on baseline electrocardiograms interpreted at each participating site, according to conventional criteria (QRS duration ≥120ms with typical LBBB morphology), consistent with AHA/ACCF/HRS recommendations. (10) No centralized ECG core laboratory adjudication was performed, reflecting a real-world clinical cohort. Patients with prior myocardial infarction, coronary artery bypass graft surgery, heart transplant, or other non-LBBB major cardiac conduction disorders (i.e., right bundle branch block, atrioventricular block, paced rhythm or Wolff-Parkinson-White syndrome) were excluded. We also included 932 control patients without conduction abnormalities who were propensity-matched in a 4:1 ratio by age, sex, and cardiovascular risk factors. The REFINE PET registry was approved by the institutional review boards of all participating institutions.

### Clinical Variables

Demographic and clinical information collected at the time of imaging included age, sex, and race; cardiovascular risk factors (hypertension, diabetes, dyslipidemia, smoking status, peripheral vascular disease); medications that could influence ischemic assessment, including beta-blockers, calcium channel blockers, nitrates, digitalis, and antiarrhythmic agents; history of percutaneous coronary intervention (PCI) or stenting; body mass index (BMI); estimated glomerular filtration rate; and hemodynamic parameters (resting and peak heart rate and blood pressure). Coronary artery calcium (CAC) score was assessed using the Agatston method, calculated automatically with deep learning algorithms (11).

### PET/CT Imaging Protocol and Quantification

All patients underwent positron emission tomography and computed tomography myocardial perfusion imaging (PET/CT MPI) using rubidium-82 (n = 953) or N-13 ammonia (n = 212) tracers, according to standard protocols. All gated PET studies met standard quality criteria for clinical interpretation. Studies with non-diagnostic gating quality were excluded per routine clinical practice. Atrial fibrillation or ventricular ectopy were not systematically excluded, reflecting real-world imaging conditions. All quantitative analyses were conducted at the centralized core laboratory for REFINE PET (Cedars-Sinai Medical Center) using validated software (QPET, Cedars-Sinai Medical Center, Los Angeles, CA) (12,13).

#### LV Remodeling and Regional and Global Systolic Function

Gated PET-derived LV volumes and ejection fraction were used to characterize patterns of left ventricular remodeling and systolic function across study groups within the same imaging modality. Indexed end-diastolic (iEDV) and end-systolic volumes (iESV) (mL/m²) were measured, and left ventricular ejection fraction (LVEF) (%) was calculated. Motion (0–5) and thickening (0–3) scoring procedures are described in the **Supplemental Methods** (13).

#### LV Mechanical Dyssynchrony

Phase entropy (%) was quantified on gated stress and rest images using Cardiac Suite (Cedars-Sinai Medical Center, Los Angeles, CA), as previously described (14–17), to assess LV mechanical dyssynchrony. In PET phase analysis, entropy summarizes the dispersion of regional contraction timing across the LV. Compared with phase standard deviation or bandwidth, entropy captures global temporal disorder of LV contraction and is less influenced by histogram shape, making it particularly suitable for complex dyssynchrony patterns such as LBBB. Entropy is derived from the phase histogram generated by first-harmonic Fourier analysis of gated LV data and increases as the phase distribution becomes broader; higher values therefore denote greater heterogeneity and mechanical dyssynchrony. Stress phase entropy was used as the primary dyssynchrony metric given its established incremental prognostic value (16,18).

#### Myocardial Blood Flow, Flow Reserve, Coronary Vascular Resistance

Global and regional stress/rest myocardial blood flow (MBF, mL/min/g) were quantified from early dynamic acquisitions with QPET software. Myocardial flow reserve (MFR) was calculated as stress MBF/rest MBF, and coronary vascular resistance (CVR) as mean arterial pressure/MBF (mmHg/mL·min⁻¹·g⁻¹)(12).

#### Myocardial Blood Flow Heterogeneity

Regional MBF was assessed in five LV regions (septal, lateral, anterior, inferior, apex), matching the approach for motion and thickening. Septal-to-lateral ratios for MBF, MFR, and CVR were calculated to characterize regional flow heterogeneity in relation to LBBB presence.

### Clinical Outcomes

We collected data on all-cause mortality and heart failure (HF) admissions as recorded in the REFINE PET registry at each participating site. Hospitalization for HF was defined by the primary discharge diagnosis, determined through previously validated International Classification of Diseases (ICD) codes. All-cause mortality and HF events were verified locally by site investigators through review of hospital electronic medical records or patient contact, following the adjudication procedures of the REFINE PET registry (9). For each event, the number of days from the PET scan to the outcome occurrence was documented.

### Statistical Analysis

To enhance comparability and reduce confounding, we matched patients with LBBB to controls 4:1 using nearest-neighbor propensity scores derived from logistic regression on age, sex, BMI, hypertension, and diabetes, with a caliper width of 0.1 of the standard deviation (SD) of the logit. Categorical variables are reported as counts (percentages) and compared with chi-square or Fisher’s exact tests. Continuous variables are presented as mean ± SD and compared with Welch’s t test. Mean differences with 95% confidence intervals (CI) quantify LBBB–control differences.

Pearson correlation coefficients were computed to assess the relationships between stress phase entropy and LV remodeling parameters (iEDV, iESV, and LVEF), as well as between stress phase entropy and the stress septal-to-lateral MBF ratio (SLR). Additionally, correlations were evaluated between the stress SLR, stress global MBF, stress global CVR, and global MFR with iEDV, iESV, and LVEF.

To examine mechanical dyssynchrony, we stratified controls and patients with LBBB into three stress phase entropy groups using cohort-wide 25th and 75th percentile cutoffs (≤25th, 25th–75th, ≥75th). For each group, we summarized phase entropy (mean, SD, minimum, maximum) and mean ± SD for iEDV, iESV, LVEF, and the stress SLR, and compared event rates (death and heart failure admissions) across strata.

Univariate linear regression analyses were used to identify PET-derived predictors of iEDV, iESV, and LVEF. Multivariable linear regression models, adjusted for age, sex, BMI, diabetes, hypertension, and CAC score, were used to test five PET MPI variables: stress phase entropy (dyssynchrony index), stress global MBF, stress global CVR, global MFR, and the stress SLR. An interaction term between LBBB status and stress SLR was included to assess effect modification on left ventricular remodeling and function. Multicollinearity among PET-derived predictors included in the multivariable models was assessed using variance inflation factors (VIF), with no evidence of problematic collinearity observed.

For survival analyses, the pooled cohort was analyzed using Cox proportional hazards models for a composite endpoint of all-cause mortality and/or heart failure hospitalization. Models were adjusted for age, sex, BMI, hypertension, diabetes, smoking status, prior PCI/stent, peripheral vascular disease, and CAC score. Predictors included LBBB status, resting LVEF, stress phase entropy, stress global MBF, stress SLR, global MFR, and stress global CVR. The proportional hazards assumption was evaluated using Schoenfeld residuals, and no violations were observed, as confirmed by visual inspection of the residual plots.

Statistical significance was set at p<0.05 (two-tailed). Propensity score and survival analyses were conducted in R (R Core Team, Vienna, Austria) using RStudio; all other analyses were performed in Stata 18.0 (StataCorp, College Station, TX).

## RESULTS

### Characteristics of the Study Cohort

The clinical characteristics of the study cohort are summarized in **Table 1**. After propensity matching, age, BMI, sex, and cardiovascular risk factors were balanced between patients with and without LBBB, with only minor differences in medication use and CAC scores. No clinically significant differences were observed between groups in mean heart rate or blood pressure at rest or during stress.

**Table 1.**
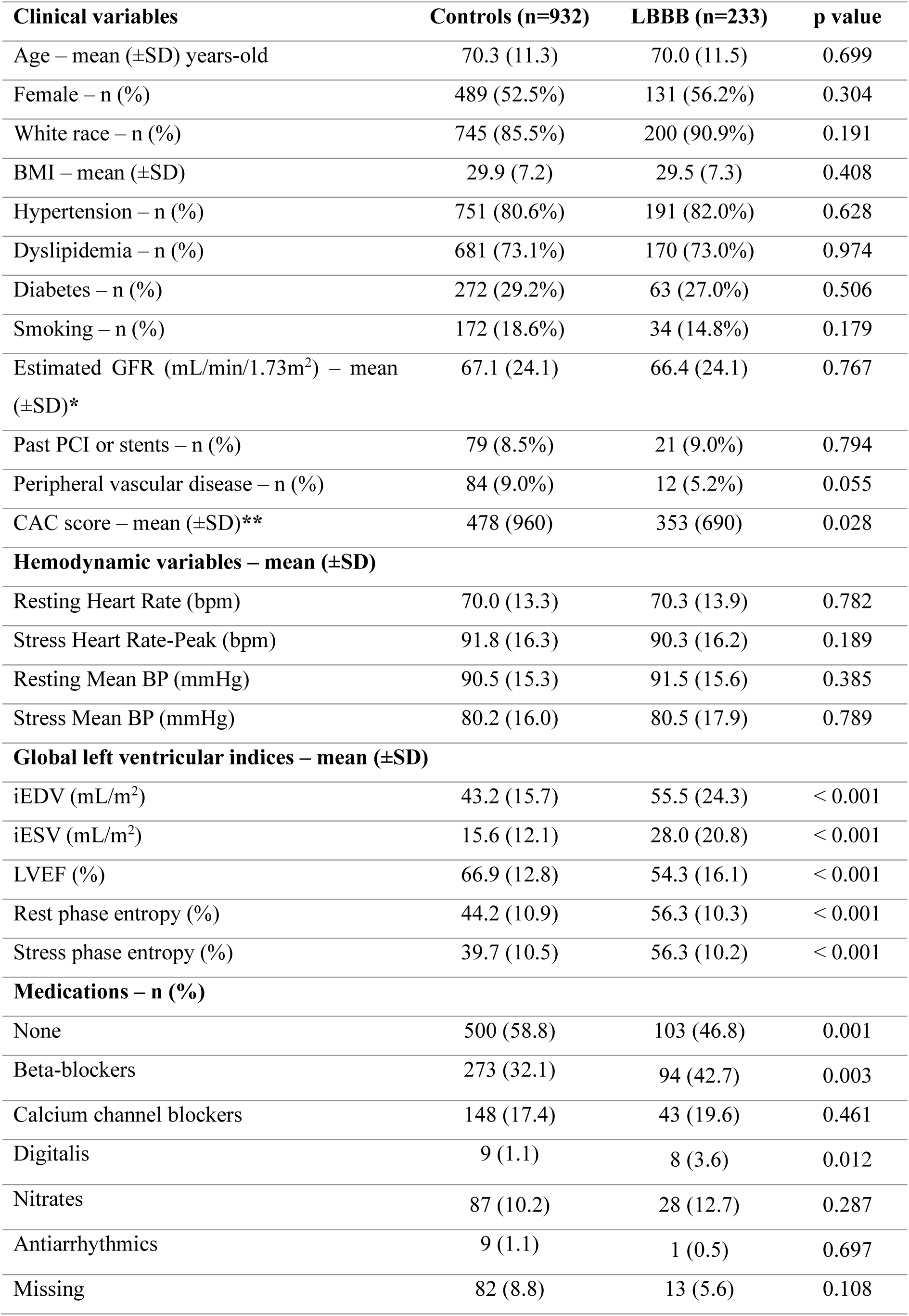
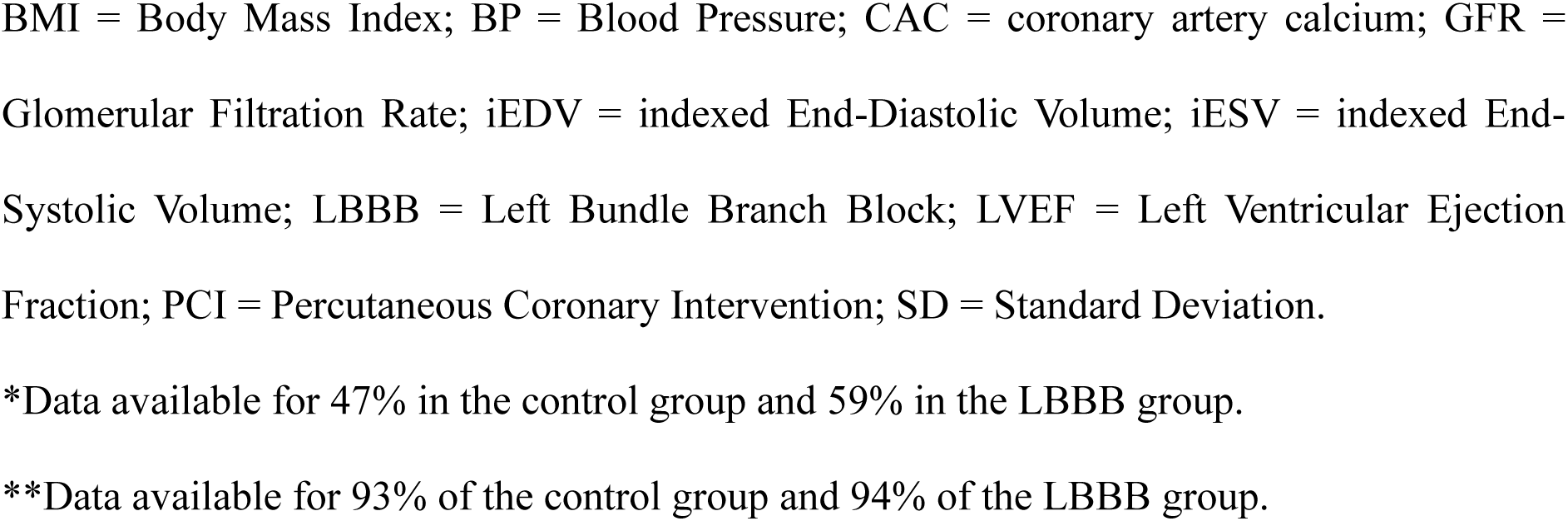
Clinical characteristics of the two groups.

| **Clinical variables** | **Controls (n=932)** | **LBBB (n=233)** | **p value** |
| --- | --- | --- | --- |
| Age – mean (±SD) years-old | 70.3 (11.3) | 70.0 (11.5) | 0.699 |
| Female – n (%) | 489 (52.5%) | 131 (56.2%) | 0.304 |
| White race – n (%) | 745 (85.5%) | 200 (90.9%) | 0.191 |
| BMI – mean (±SD) | 29.9 (7.2) | 29.5 (7.3) | 0.408 |
| Hypertension – n (%) | 751 (80.6%) | 191 (82.0%) | 0.628 |
| Dyslipidemia – n (%) | 681 (73.1%) | 170 (73.0%) | 0.974 |
| Diabetes – n (%) | 272 (29.2%) | 63 (27.0%) | 0.506 |
| Smoking – n (%) | 172 (18.6%) | 34 (14.8%) | 0.179 |
| Estimated GFR (mL/min/1.73m<sup>2</sup>) – mean (±SD)* | 67.1 (24.1) | 66.4 (24.1) | 0.767 |
| Past PCI or stents – n (%) | 79 (8.5%) | 21 (9.0%) | 0.794 |
| Peripheral vascular disease – n (%) | 84 (9.0%) | 12 (5.2%) | 0.055 |
| CAC score – mean (±SD)** | 478 (960) | 353 (690) | 0.028 |
| **Hemodynamic variables – mean (±SD)** |  |  |  |
| Resting Heart Rate (bpm) | 70.0 (13.3) | 70.3 (13.9) | 0.782 |
| Stress Heart Rate-Peak (bpm) | 91.8 (16.3) | 90.3 (16.2) | 0.189 |
| Resting Mean BP (mmHg) | 90.5 (15.3) | 91.5 (15.6) | 0.385 |
| Stress Mean BP (mmHg) | 80.2 (16.0) | 80.5 (17.9) | 0.789 |
| **Global left ventricular indices – mean (±SD)** |  |  |  |
| iEDV (mL/m<sup>2</sup>) | 43.2 (15.7) | 55.5 (24.3) | < 0.001 |
| iESV (mL/m<sup>2</sup>) | 15.6 (12.1) | 28.0 (20.8) | < 0.001 |
| LVEF (%) | 66.9 (12.8) | 54.3 (16.1) | < 0.001 |
| Rest phase entropy (%) | 44.2 (10.9) | 56.3 (10.3) | < 0.001 |
| Stress phase entropy (%) | 39.7 (10.5) | 56.3 (10.2) | < 0.001 |
| **Medications – n (%)** |  |  |  |
| None | 500 (58.8) | 103 (46.8) | 0.001 |
| Beta-blockers | 273 (32.1) | 94 (42.7) | 0.003 |
| Calcium channel blockers | 148 (17.4) | 43 (19.6) | 0.461 |
| Digitalis | 9 (1.1) | 8 (3.6) | 0.012 |
| Nitrates | 87 (10.2) | 28 (12.7) | 0.287 |
| Antiarrhythmics | 9 (1.1) | 1 (0.5) | 0.697 |
| Missing | 82 (8.8) | 13 (5.6) | 0.108 |

BMI = Body Mass Index; BP = Blood Pressure; CAC = coronary artery calcium; GFR = Glomerular Filtration Rate; iEDV = indexed End-Diastolic Volume; iESV = indexed End-Systolic Volume; LBBB = Left Bundle Branch Block; LVEF = Left Ventricular Ejection Fraction; PCI = Percutaneous Coronary Intervention; SD = Standard Deviation.
\*Data available for 47% in the control group and 59% in the LBBB group.
\*\*Data available for 93% of the control group and 94% of the LBBB group.

### LV Remodeling, Function, and Mechanical Dyssynchrony

Compared with controls, patients with LBBB demonstrated significantly larger iEDV and iESV, as well as lower LVEF. In the overall study population, 186 patients (16%) had an LVEF <50%, including 96 of 932 controls (10%) and 90 of 233 patients with LBBB (39%). As expected, patients with LBBB also exhibited significantly greater mechanical dyssynchrony than controls (**Table 1**). Among left ventricular regions, the septal wall and the apex showed the most pronounced functional impairment in the LBBB group **(Supplemental Table S1)**.

### Myocardial Perfusion Metrics and Coronary Vascular Resistance

At rest, global MBF (1.08 ± 0.37 vs 1.10 ± 0.39 mL/min/g, p = 0.584) and CVR (93.57 ± 34.42 vs 92.29 ± 36.84 mmHg/mL/min/g, p = 0.616) were similar in patients with LBBB and controls, respectively. During stress, global MBF was lower (2.43 ± 0.77 vs 2.59 ± 0.80 mL/min/g, p = 0.004) and CVR higher (36.50 ± 15.12 vs 34.05 ± 13.64 mmHg/mL/min/g, p = 0.025) in LBBB patients than in controls. Consequently, global MFR was lower in LBBB than in controls (2.41 ± 0.75 vs 2.55 ± 0.84 mL/min/g, p = 0.013).

To highlight regional differences in MBF and CVR we created a septal-to-lateral myocardial blood flow ratio (SLR). Compared to controls, patients with LBBB had lower rest (0.87 ± 0.24 vs 0.93 ± 0.16, p <0.001) and stress (0.95 ± 0.18 vs 1.00 ± 0.17, p <0.001) SLR, and higher rest (1.21 ± 0.33 vs 1.10 ± 0.22, p < 0.001) and stress (1.09 ± 0.24 vs 1.03 ± 0.19, respectively, p < 0.001) septal-to-lateral CVR ratios. A complete description of regional differences in MBF, MFR, and CVR is included in **Supplemental Table S2**.

### Interplay between Mechanical Dyssynchrony, Myocardial Perfusion Heterogeneity, and LV Remodeling

Figure 1 shows the distribution of mechanical dyssynchrony as phase entropy percentile-based categories of severity (≤25th, 25th–75th, and ≥75th; with higher percentile category representing more severe dyssynchrony) in LBBB and control patients. Most patients with LBBB (72.4%) were classified in the highest percentile category, whereas only 13.1% of controls fell into this category. Conversely, only 2.6% of patients with LBBB but 30.7% of controls were in the lowest category. Of note, 25% of patients with LBBB were in the intermediate category (25th–75th percentile), compared with 56% of controls.

**Figure 1.**
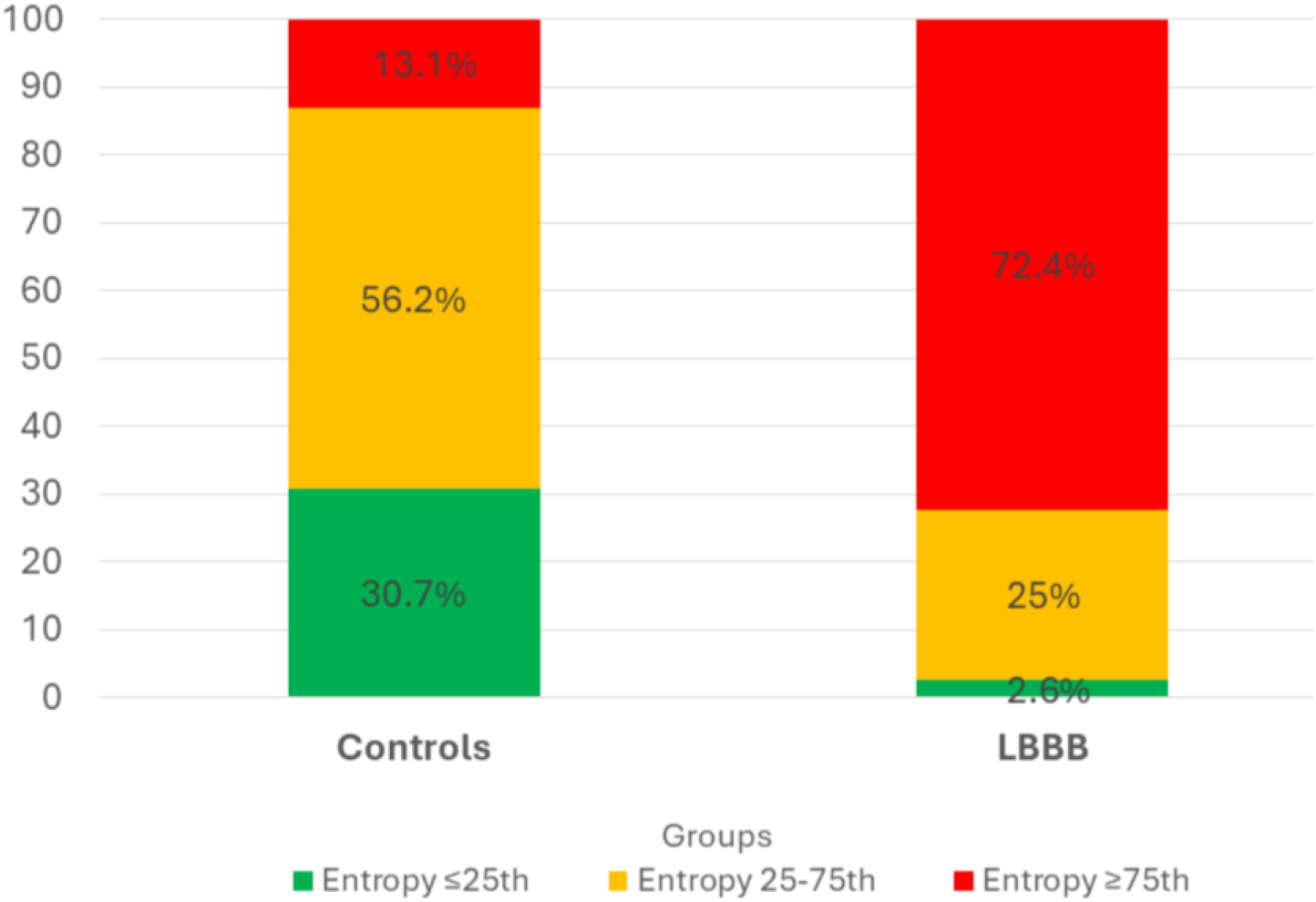
Distribution of left ventricular (LV) stress phase entropy values by percentile (<25th, 25th–75th, and >75th) in the control and left bundle branch block (LBBB) groups. Percentile categories were defined based on the distribution of **stress phase entropy values**, which are expressed as percentages. Entropy thresholds corresponding to the <25th, 25th–75th, and >75th percentiles were **<34%, 34–52%, and >52%**, respectively, and were applied uniformly to both control subjects and patients with LBBB. A higher proportion of patients with LBBB exhibited entropy values above the 75th percentile compared with controls (72.4% vs. 13.1%), whereas most controls fell within the 25th–75th percentile range. Higher stress phase entropy reflects greater mechanical dyssynchrony (i.e., worse LV synchrony). Values within each bar represent the percentage of individuals in each entropy percentile category.

**Table 2** shows the independent PET MPI predictors of adverse LV remodeling and dysfunction in the combined cohort (with LBBB entered as a covariate): stress phase entropy, stress SLR, global stress CVR, and global stress MBF remained in the final model, whereas global MFR was not significant. We also observed a significant LBBB × stress SLR interaction for iEDV, iESV, and LVEF, indicating that the adverse association of a reduced SLR with remodeling and dysfunction is amplified in LBBB. As shown in Figure 2, the increases in iEDV and iESV per unit decrease in SLR are approximately three to four times greater in LBBB than in controls, with a correspondingly steeper reduction in LVEF.

**Figure 2.**
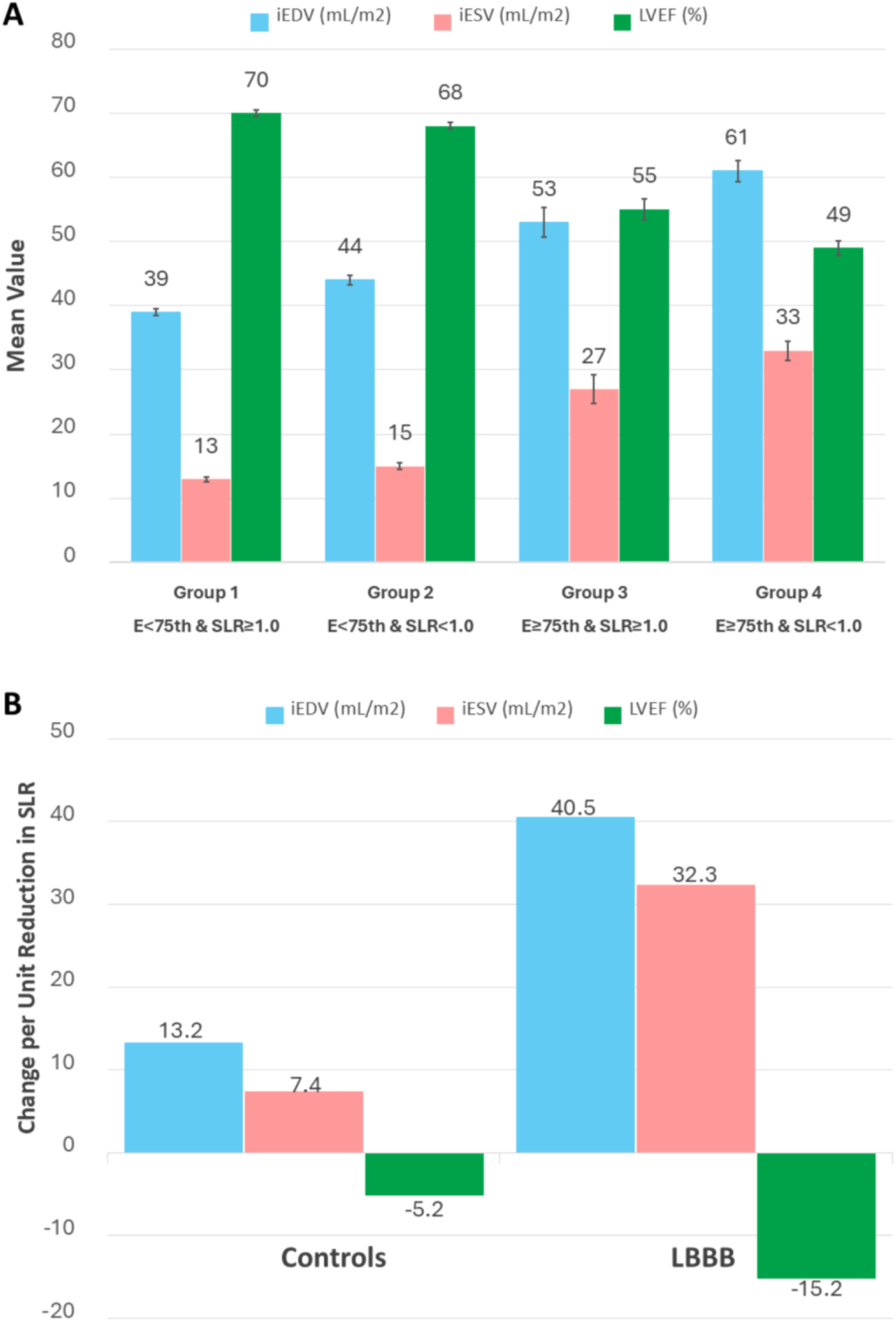
PET Phenotypes, LV Remodeling, and Impact of Perfusion Heterogeneity. **(A) LV Remodeling and Function Across Risk Groups.** Patients (n = 1161) were classified into four groups according to stress phase entropy (<75th vs. ≥75th percentile) and the stress septal-to-lateral myocardial blood-flow ratio (SLR; ≥1.0 vs. <1.0). This classification reflects increasing mechanical dyssynchrony and myocardial blood-flow heterogeneity. Progression from Groups 1→4 (low to high risk) was associated with larger LV volumes (iEDV, iESV) and lower LVEF, indicating additive adverse effects of dyssynchrony and perfusion heterogeneity. All differences across groups were statistically significant at p < 0.05. **(B) Adverse Effect of Reduced SLR by Group.** A reduced SLR had a stronger adverse effect in LBBB patients compared with controls, amplifying LV remodeling and dysfunction by approximately 3–4× (interaction). iEDV = indexed end-diastolic volume; iESV = indexed end-systolic volume; LVEF = left ventricular ejection fraction; LBBB = left bundle branch block; SLR = septal-to-lateral myocardial blood-flow ratio.

**Table 2.** Factors associated with left ventricular remodeling (iEDV and iESV) and function (LVEF) in the total sample, with LBBB included as a covariate.

| Associations | iEDV (n= 1074)<br>Coefficient<br>(95%IC)<br>p value |  | iESV (n= 1074)<br>Coefficient<br>(95%IC)<br>p value |  | LVEF (n= 1072)<br>Coefficient<br>(95%IC)<br>p value |  |
| --- | --- | --- | --- | --- | --- | --- |
|  | Univariate | Multivariate | Univariate | Multivariate | Univariate | Multivariate |
| LBBB | 47.67<br>(34.28, 61.07)<br>< 0.001 | 30.73<br>(18.18, 43.28)<br>< 0.001 | 45.16<br>(34.30, 56.01)<br>< 0.001 | 28.10<br>(18.15, 38.04)<br>< 0.001 | -32.26<br>(-42.79, -21.73)<br>< 0.001 | -12.53<br>(-21.40, -3.65)<br>0.006 |
| SLR | -17.78<br>(-24.31, -11.24)<br>< 0.001 | -13.22<br>(-19.28, -7.16)<br>< 0.001 | -11.21<br>(-16.50, -5.91)<br>< 0.001 | -7.44<br>(12.24, -2.63)<br>0.002 | 8.94<br>(3.80, 14.07)<br>0.001 | 5.22<br>(0.93, 9.51)<br>0.017 |
| LBBB*SLR | -37.71<br>(-51.35, -24.07)<br>< 0.001 | -27.27<br>(-39.88, -14.66)<br>< 0.001 | -34.75<br>(-45.81, -23.70)<br>< 0.001 | -24.82<br>(-34.82, -14.82)<br>< 0.001 | 21.05<br>(10.33, 31.77)<br>< 0.001 | 10.01<br>(1.08, 18.93)<br>0.028 |
| Stress Entropy | 0.57<br>(0.49, 0.65)<br>< 0.001 | 0.38<br>(0.29, 0.47)<br>< 0.001 | 0.59<br>(0.53, 0.65)<br>< 0.001 | 0.44<br>(0.37, 0.51)<br>< 0.001 | -0.66<br>(-0.72, -0.61)<br>< 0.001 | -0.56<br>(-0.62; -0.49)<br>< 0.001 |
| Global MBF | -7.88<br>(-9.13, -6.64)<br>< 0.001 | -2.05<br>(-3.78, -0.32)<br>0.02 | -6.37<br>(-7.40, 5.34)<br>< 0.001 | -1.64<br>(-3.01,-0.27)<br>0.019 | 6.55<br>(5.58, 7.53)<br>< 0.001 | 1.87<br>(0.65; 3.10)<br>0.003 |
| Global CVR | 0.50<br>(0.43, 0.57)<br>< 0.001 | 0.23<br>(0.13, 0.33)<br>< 0.001 | 0.39<br>(0.33, 0.45)<br>< 0.001 | 0.15<br>(0.07, 0.23)<br>< 0.001 | -0.37<br>(-0.43, -0.32)<br>< 0.001 | -0.11<br>(-0.18, -0.04)<br>0.002 |

CVR = Coronary Vascular Resistance during stress; iEDV = indexed End-Diastolic Volume; iESV = indexed End-Systolic Volume; LBBB = Left Bundle Branch Block; LBBB\*SLR = interaction between Left Bundle Branch Block and Septal-to-Lateral Myocardial Blood Flow ratio during stress; LVEF = Left Ventricular Ejection Fraction; MBF = Myocardial Blood Flow; SLR = Septal-to-Lateral Myocardial Blood Flow ratio during stress.

To understand the substrate for this effect modification, we examined how dyssynchrony and perfusion heterogeneity cluster across groups (**Table 3**). Higher stress phase entropy—reflecting advanced mechanical dyssynchrony—was associated with stepwise increases in LV volumes and lower LVEF in both controls and patients with LBBB, with a more pronounced gradient in LBBB. Importantly, only patients with LBBB with advanced dyssynchrony (≥75th percentile of entropy) exhibited a mean stress SLR below 1.0 (0.92 ± 0.18), consistent with reduced relative septal perfusion; all other subgroups maintained mean SLR ≥1.0, suggesting preserved septal–lateral balance. Correlations between stress phase entropy, stress SLR, and LV volumes/function are provided in **Supplemental Figure S1**.

**Table 3.** Distribution of left ventricular remodeling, function, myocardial perfusion heterogeneity, and clinical outcomes across stress phase entropy percentiles in controls and LBBB patients.

|  | Controls (N = 929) |  |  |  | LBBB (N = 232) |  |  |  |
| --- | --- | --- | --- | --- | --- | --- | --- | --- |
| Variables - mean ± SD | ≤25<sup>th</sup><br>n = 285 | 25-75<sup>th</sup><br>n = 522 | ≥75<sup>th</sup><br>n = 122 | p<br>Value | ≤25<sup>th</sup><br>n = 6 | 25-75<sup>th</sup><br>n = 58 | ≥75<sup>th</sup><br>n = 168 | p<br>Value |
| Phase entropy (%)<br>(Lower and Upper) | 28 ± 4<br>(9-37) | 42 ± 5<br>(34-52) | 59 ± 6<br>(52-79) | - | 26 ± 4<br>(21-31) | 45 ± 5<br>(34-52) | 61 ± 6<br>(52-77) | - |
| LVEF(%) | 73 ± 9 | 67 ± 10 | 52 ± 16 | <0.001 | 68 ± 16* | 62 ± 14* | 51 ± 16 | <0.001 |
| iEDV (mL/m<sup>2</sup>) | 39 ± 13 | 43 ± 4 | 55 ± 22 | <0.001 | 32 ± 9* | 45 ± 22* | 60 ± 24 | <0.001 |
| iESV (mL/m<sup>2</sup>) | 11 ± 7 | 15 ± 9. | 29 ± 21 | <0.001 | 11 ± 7* | 19 ± 15* | 32 ± 22 | <0.001 |
| Stress SLR | 1.02 ±<br>0.15 | 1.00 ±<br>0.16 | 1.00 ±<br>0.22 | 0.218 | 1.08 ±<br>0.09* | 1.05 ±<br>0.16* | 0.92 ±<br>0.18 | <0.001 |
| Death - n (%) | 32 (11%) | 107 (21%) | 59 (48%) | <0.001 | 1 (17%) | 14 (24%) | 50 (30%) | 0.586 |
| HF- n (%) | 6 (2%) | 29 (6%) | 19 (16%) | <0.001 | 0 (0) | 5 (9%) | 21 (13%) | 0.491 |

The table compares ventricular volumes, function, myocardial perfusion heterogeneity, and outcomes (death and heart failure admissions) across these groups. The stress septal-to-lateral MBF ratio (SLR) underscores the link between mechanical dyssynchrony and perfusion heterogeneity in LBBB.
HF = Heart failure hospitalizations; iEDV = Indexed end-diastolic volume; iESV = Indexed end-systolic volume; LBBB = Left bundle branch block; LVEF = Left ventricular ejection fraction; MBF = Myocardial blood flow; Phase entropy = Phase measure of LV mechanical dyssynchrony during stress; SLR = Septal-to-lateral (myocardial blood flow) ratio; SD = Standard deviation.
*\*The difference between the ≤25<sup>th</sup> and 25–75<sup>th</sup> groups was not statistically significant.*

Accordingly, we further explored the interplay between perfusion heterogeneity and dyssynchrony severity to gauge LV remodeling severity by stratifying all patients according to both stress SLR (<1.0 vs ≥1.0) and phase entropy (<75th vs ≥75th percentile). This approach identified four distinct phenotypic groups that exhibited a progressive pattern of greater LV remodeling and lower LV function, as shown in Figure 2 and the **Central Illustration** (additional details in **Supplemental Table S3**).

### Survival Analysis

During a mean follow-up of 5.2 years, 65 patients (28%) with LBBB and 198 controls (21%) died (p = 0.037). HF hospitalization occurred in 26 patients (11%) with LBBB and 54 controls (6%) (p = 0.006).

In Cox regression analysis adjusting for age, sex, BMI, hypertension, diabetes, smoking, CAC score, prior PCI, and peripheral vascular disease, stress phase entropy (adjusted HR: 1.02, p = 0.011), global MFR (adjusted HR: 0.62, p < 0.001), and resting LVEF (adjusted HR: 0.97, p < 0.001) emerged as independent predictors of the composite endpoint of all-cause mortality and/or heart failure admission. After multivariable adjustment, LBBB was not significantly associated with the composite endpoint of all-cause mortality or heart failure hospitalization (**Table 4**).

**Table 4.**
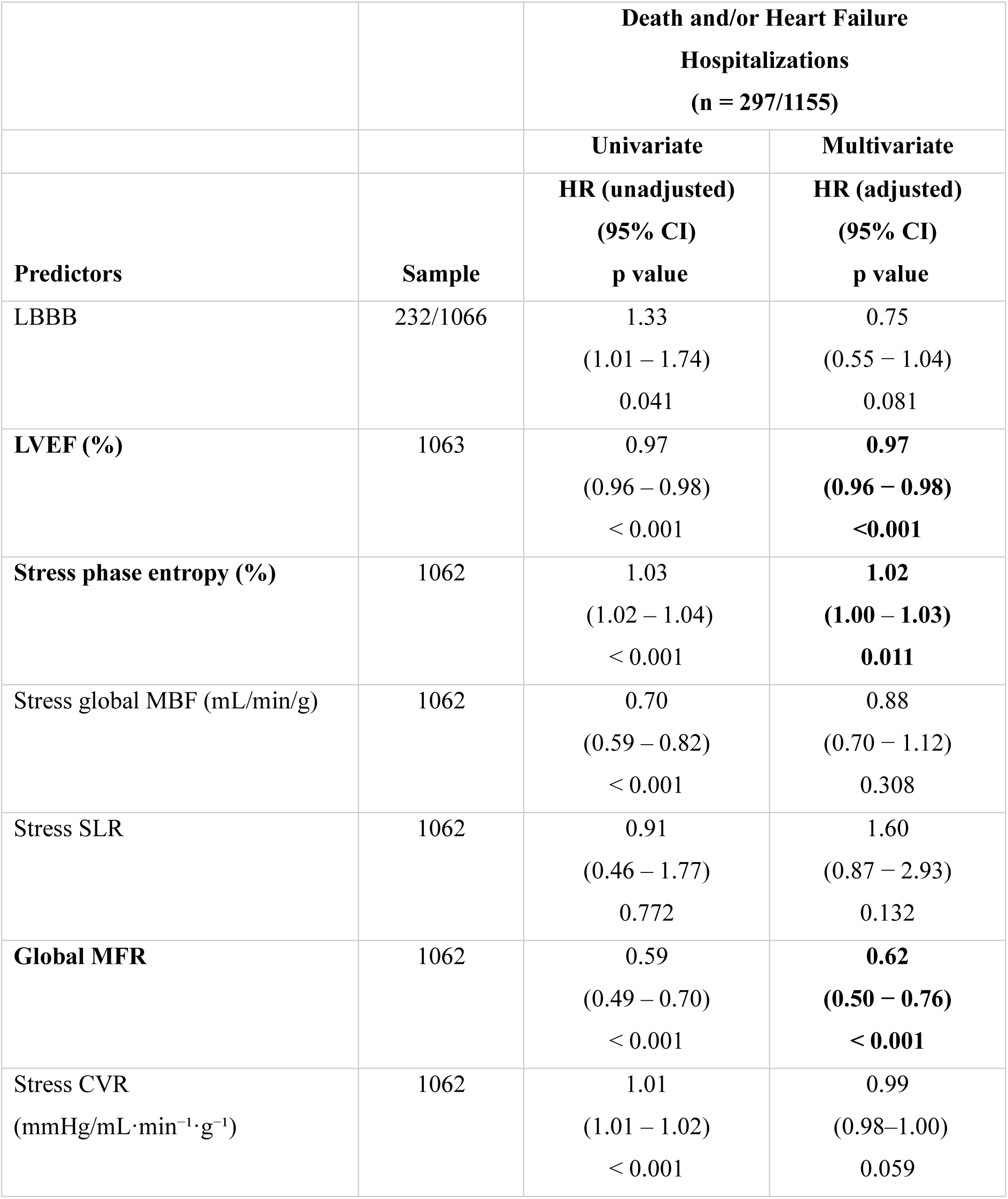
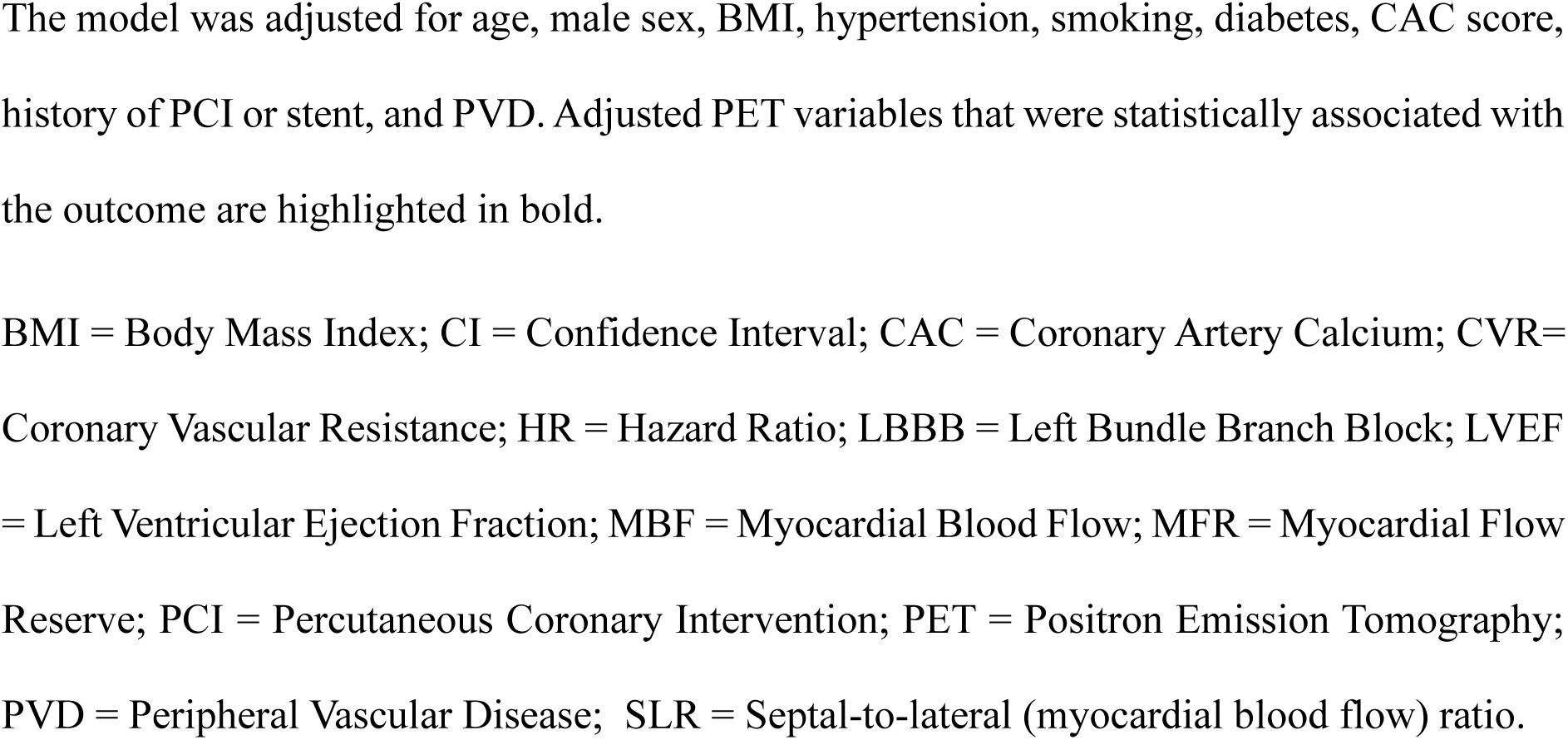
Factors associated with death and heart failure hospitalizations in the total sample, including LBBB as a covariate.

| Predictors | Sample | Death and/or Heart Failure Hospitalizations<br>(n = 297/1155) |  |
| --- | --- | --- | --- |
|  |  | Univariate | Multivariate |
|  |  | HR (unadjusted)<br>(95% CI)<br>p value | HR (adjusted)<br>(95% CI)<br>p value |
| LBBB | 232/1066 | 1.33<br>(1.01 – 1.74)<br>0.041 | 0.75<br>(0.55 – 1.04)<br>0.081 |
| LVEF (%) | 1063 | 0.97<br>(0.96 – 0.98)<br>< 0.001 | **0.97**<br>**(0.96 – 0.98)**<br>**<0.001** |
| Stress phase entropy (%) | 1062 | 1.03<br>(1.02 – 1.04)<br>< 0.001 | **1.02**<br>**(1.00 – 1.03)**<br>**0.011** |
| Stress global MBF (mL/min/g) | 1062 | 0.70<br>(0.59 – 0.82)<br>< 0.001 | 0.88<br>(0.70 – 1.12)<br>0.308 |
| Stress SLR | 1062 | 0.91<br>(0.46 – 1.77)<br>0.772 | 1.60<br>(0.87 – 2.93)<br>0.132 |
| Global MFR | 1062 | 0.59<br>(0.49 – 0.70)<br>< 0.001 | **0.62**<br>**(0.50 – 0.76)**<br>**< 0.001** |
| Stress CVR<br>(mmHg/mL·min<sup>-1</sup>·g<sup>-1</sup>) | 1062 | 1.01<br>(1.01 – 1.02)<br>< 0.001 | 0.99<br>(0.98–1.00)<br>0.059 |

The model was adjusted for age, male sex, BMI, hypertension, smoking, diabetes, CAC score, history of PCI or stent, and PVD. Adjusted PET variables that were statistically associated with the outcome are highlighted in bold.
BMI = Body Mass Index; CI = Confidence Interval; CAC = Coronary Artery Calcium; CVR= Coronary Vascular Resistance; HR = Hazard Ratio; LBBB = Left Bundle Branch Block; LVEF = Left Ventricular Ejection Fraction; MBF = Myocardial Blood Flow; MFR = Myocardial Flow Reserve; PCI = Percutaneous Coronary Intervention; PET = Positron Emission Tomography; PVD = Peripheral Vascular Disease; SLR = Septal-to-lateral (myocardial blood flow) ratio.

## DISCUSSION

In this definitive multicenter PET study—the largest to date assessing patients with LBBB using PET MPI alongside a well-matched control group without conduction disturbances—we provide robust evidence on the impact of LBBB on coronary vascular function. Unlike prior studies(5,8,14,15), which were generally limited by small sample sizes, single-center designs, absence of appropriate controls, or a focus on advanced LV dysfunction, our work integrates absolute and relative perfusion metrics with mechanical assessment. We found that LBBB is characterized by advanced mechanical dyssynchrony and a consistent septal-to-lateral MBF imbalance. Both abnormalities independently and synergistically predicted adverse LV remodeling and systolic dysfunction, and their interaction identified distinct phenotypes with varying risk of adverse LV remodeling.

This result may explain why some LBBB patients maintain preserved LV function, while others progress to heart failure and ventricular dilation (16). LBBB causes LV electrical and mechanical dyssynchrony, leading to abnormal myocardial contraction. This abnormal activation pattern, over time, can result in adverse LV remodeling increasing the risk of heart failure and sudden cardiac death. However, in the absence of underlying cardiovascular disease, there is heterogeneity in the frequency and timing of progression to LV systolic dysfunction with up to two thirds of patients not progressing to LV dysfunction (17). The underlying mechanisms for this heterogeneity are not well understood, although an interplay between myocardial hypoperfusion and fibrosis have been implicated (18).

As expected, in our study most patients with LBBB demonstrated significant mechanical dyssynchrony, reflected by markedly elevated phase entropy. Interestingly, 13% of controls without LBBB also exhibited significant dyssynchrony, which was associated with remodeling, impaired LV function, and higher mortality. This finding confirms those of prior studies (19,20) and underscores that mechanical dyssynchrony is not exclusive to LBBB, and that LBBB itself is heterogeneous—some patients maintain preserved synchrony and function, while others progress to cardiomyopathy. The SLR did not independently predict outcomes but retained value as an early marker of regional dysfunction (21). However, LVEF, stress phase entropy, and global MFR were prognostic indicators for HF hospitalization and mortality (22–24).

The classic explanation for reduced septal perfusion in LBBB; that is, lower metabolic demand from early-activated myocardium, does not fully capture the complexity we observed (6,8). We found that both phase entropy and the SLR were independent predictors of adverse remodeling in LBBB and controls, but their associations were significantly stronger in LBBB. This supports the notion that conduction disturbance amplifies the adverse structural effects of microvascular heterogeneity. The data suggests that the SLR likely reflects more than simply differences in oxygen demand, and that it may provide a quantitative marker that integrates the mechanical and coronary vascular consequences of conduction delay. Our data extend prior observations (25,26) by demonstrating that LBBB induces not only septal hypoperfusion and metabolic downregulation (14,15,27), but it is also associated with reduced stress MBF and flow reserve in the lateral wall relative to subjects without conduction abnormalities.

Exactly how the interaction between mechanical dyssynchrony and perfusion heterogeneity leads to adverse LV remodeling cannot be fully determined from this study, but our data suggest two distinct mechanistic pathways. In patients with LBBB, electrical dyssynchrony appears to be the primary driver of remodeling. The delayed activation pattern chronically redistributes workload and intramyocardial compressive forces, creating a persistent septal–lateral supply–demand mismatch. This regional imbalance increases septal vascular resistance and reduces stress MBF, reflecting functional microvascular dysfunction.

Over time, the sustained underperfusion of the septum promotes fibrosis and progressive LV remodeling (28). In contrast, among controls without conduction defects, mechanical dyssynchrony likely represents a secondary phenomenon, arising from underlying myocardial or fibrotic processes. In these patients, the relationship between a reduced SLR and LV remodeling was mechanistically similar but approximately three to four times weaker than in LBBB. This pattern suggests that primary electrical dyssynchrony in LBBB can initiate a cardiomyopathic process, whereas secondary dyssynchrony in structurally abnormal hearts may amplify pre-existing myocardial disease.

Together, these findings support a bidirectional model in which electrical conduction delay and myocardial injury reinforce each other through microvascular dysfunction and regional flow imbalance. This feedback loop may sustain a cycle of mechanical inefficiency, progressive fibrosis, and LV dilation—linking conduction disturbance to structural heart disease.

In experimental models of LBBB, regional ^99m^Tc-sestamibi uptake on SPECT imaging correlated with myocardial work and the degree of endocardial collagen content (18). As compensation for myocardial injury, fibroblasts are activated and modulate the composition of the interstitial extracellular matrix, which in turn may lead to microvascular dysfunction, leading to a vicious cycle of myocardial dysfunction, perfusion heterogeneity, hypoperfusion, and fibrosis. Therefore, our results confirm that LBBB is not merely an electrical phenomenon—it initiates early mechanical and microvascular changes that can progress to global dysfunction (3). Advanced imaging identifies high-risk phenotypes not evident on ECG alone, mirroring findings from the cardiac resynchronization therapy literature, where restoring synchrony can, in some but not all cases, normalize perfusion and reverse remodeling (29,30).

## Limitations

Several limitations merit consideration. First, residual confounding cannot be excluded despite rigorous matching. Second, the cross-sectional design precludes causal inference; prospective studies are needed. Third, subtle coronary disease may still influence perfusion despite exclusion of overt ischemia, though the mechanistic links with dyssynchrony support a conduction-related origin. Fourth, QRS duration was not available for analysis, preventing direct comparison with phase entropy; however, prior work has shown stronger associations of phase variables with outcomes than QRS duration (31). Finally, pharmacologic stress was used uniformly, which may produce different perfusion patterns than exercise stress. However, patients with LBBB are generally recommended for pharmacologic stress in clinical practice, so this approach reflects real-world practice rather than a study limitation.

## CONCLUSION

In summary, patients with LBBB exhibited significantly lower global and regional stress MBF and MFR, alongside increased coronary resistance—particularly in the septal wall—compared to controls. Mechanical dyssynchrony and septal–lateral perfusion heterogeneity are independent and synergistic drivers of LV remodeling and dysfunction in patients with and without LBBB. Electrical delay alone is insufficient to characterize the full scope of LBBB-related myocardial changes. Integrating phase analysis with regional flow metrics provides a more comprehensive phenotyping strategy, with potential to improve risk stratification and guide timely intervention.

### Clinical implications

Compared with patients without conduction abnormalities, LBBB is associated with reduced global stress MBF, lower MFR, and higher global stress CVR, indicating microvascular dysfunction beyond conduction delay. Stress phase entropy and the stress SLR provide incremental information not captured by surface ECG.

### Translational Implications

Advanced PET phenotyping refines risk assessment in patients with and without LBBB. Integrating measures of mechanical dyssynchrony and regional perfusion heterogeneity may improve patient selection for cardiac resynchronization therapy, enable earlier identification of preclinical cardiomyopathy, and inform preventive strategies.

## Data Availability

All data produced in the present study are available upon reasonable request to the authors.

## Acknowledgements

None

## Abbreviations

CVR —: Coronary Vascular Resistance
iEDV —: Indexed End-Diastolic Volume
EF —: Ejection Fraction
iESV —: Indexed End-Systolic Volume
LBBB —: Left Bundle Branch Block
LV —: Left Ventricle; Left Ventricular
LVEF —: Left Ventricular Ejection Fraction
MBF —: Myocardial Blood Flow
MFR —: Myocardial Flow Reserve
SLR —: Septal-to-Lateral (myocardial blood flow) Ratio

## References

1. Fahy GJ, Pinski SL, Miller DP, McCabe N, Pye C, Walsh MJ, et al. Natural history of isolated bundle branch block. Am J Cardiol. junho de 1996;77(14):1185–90.

2. Surkova E, Badano LP, Bellu R, Aruta P, Sambugaro F, Romeo G, et al. Left bundle branch block: from cardiac mechanics to clinical and diagnostic challenges. EP Europace. 1o de agosto de 2017;19(8):1251–71.

3. Huizar JF, Kaszala K, Tan A, Koneru J, Mankad P, Kron J, et al. Abnormal Conduction-Induced Cardiomyopathy. J Am Coll Cardiol. março de 2023;81(12):1192–200.

4. Ashraf H. Natural History and Clinical Significance of Isolated Complete Left Bundle Brunch Block without Associated Structural Heart Disease. The Anatolian Journal of Cardiology. 2020;

5. Lindner O, Vogt J, Baller D, Kammeier A, Wielepp P, Holzinger J, et al. Global and regional myocardial oxygen consumption and blood flow in severe cardiomyopathy with left bundle branch block. Eur J Heart Fail. março de 2005;7(2):225–30.

6. Vernooy K, Verbeek XAAM, Peschar M, Crijns HJGM, Arts T, Cornelussen RNM, et al. Left bundle branch block induces ventricular remodelling and functional septal hypoperfusion. Eur Heart J. 1o de janeiro de 2005;26(1):91–8.

7. Grines CL, Bashore TM, Boudoulas H, Olson S, Shafer P, Wooley CF. Functional abnormalities in isolated left bundle branch block. The effect of interventricular asynchrony. Circulation. abril de 1989;79(4):845–53.

8. Koepfli P, Wyss CA, Gaemperli O, Siegrist PT, Klainguti M, Schepis T, et al. Left bundle branch block causes relative but not absolute septal underperfusion during exercise. Eur Heart J. 2 de dezembro de 2009;30(24):2993–9.

9. Ramirez G, Lemley M, Shanbhag A, Kwiecinski J, Miller RJH, Kavanagh PB, et al. The REgistry of Flow and Perfusion Imaging for Artificial Intelligence with positron emission tomography (REFINE PET):Rationale and design. Journal of Nuclear Cardiology. outubro de 2025;52:102449.

10. Surawicz B, Childers R, Deal BJ, Gettes LS. AHA/ACCF/HRS Recommendations for the Standardization and Interpretation of the Electrocardiogram. Part III: Intraventricular Conduction Disturbances A Scientific Statement From the American Heart Association Electrocardiography and Arrhythmias Committee, Council on Clinical Cardiology; the American College of Cardiology Foundation; and the Heart Rhythm Society. Vol. 53, Journal of the American College of Cardiology. 2009. p. 976–81.

11. Pieszko K, Shanbhag A, Killekar A, Miller RJH, Lemley M, Otaki Y, et al. Deep Learning of Coronary Calcium Scores From PET/CT Attenuation Maps Accurately Predicts Adverse Cardiovascular Events. JACC Cardiovasc Imaging. maio de 2023;16(5):675–87.

12. Murthy VL, Bateman TM, Beanlands RS, Berman DS, Borges-Neto S, Chareonthaitawee P, et al. Clinical Quantification of Myocardial Blood Flow Using PET: Joint Position Paper of the SNMMI Cardiovascular Council and the ASNC. Journal of Nuclear Medicine. fevereiro de 2018;59(2):273–93.

13. Nakazato R, Berman DS, Alexanderson E, Slomka P. Myocardial perfusion imaging with PET. Imaging Med. fevereiro de 2013;5(1):35–46.

14. Neri G. Effect of biventricular pacing on metabolism and perfusion in patients affected by dilated cardiomyopathy and left bundle branch block: evaluation by positron emission tomography. Europace. janeiro de 2003;5(1):111–5.

15. Masci PG, Marinelli M, Piacenti M, Lorenzoni V, Positano V, Lombardi M, et al. Myocardial Structural, Perfusion, and Metabolic Correlates of Left Bundle Branch Block Mechanical Derangement in Patients With Dilated Cardiomyopathy. Circ Cardiovasc Imaging. julho de 2010;3(4):482–90.

16. Sharma S, Barot H V., Schwartzman AD, Ganatra S, Shah SP, Venesy DM, et al. Risk and predictors of dyssynchrony cardiomyopathy in left bundle branch block with preserved left ventricular ejection fraction. Clin Cardiol. 17 de dezembro de 2020;43(12):1494–500.

17. Sze E, Dunning A, Loring Z, Atwater BD, Chiswell K, Daubert JP, et al. Comparison of Incidence of Left Ventricular Systolic Dysfunction Among Patients With Left Bundle Branch Block Versus Those With Normal QRS Duration. Am J Cardiol. dezembro de 2017;120(11):1990–7.

18. Wang X, Ge B, Miao C, Lee C, Romero JE, Li P, et al. Beyond conduction impairment: Unveiling the profound myocardial injury in left bundle branch block. Heart Rhythm. agosto de 2024;21(8):1370–9.

19. Emkanjoo Z, Esmaeilzadeh M, Mohammad Hadi N, Alizadeh A, Tayyebi M, Sadrameli MA. Frequency of inter- and intraventricular dyssynchrony in patients with heart failure according to QRS width. Europace. 3 de outubro de 2007;9(12):1171–6.

20. van Bommel RJ, Tanaka H, Delgado V, Bertini M, Borleffs CJW, Ajmone Marsan N, et al. Association of intraventricular mechanical dyssynchrony with response to cardiac resynchronization therapy in heart failure patients with a narrow QRS complex. Eur Heart J. 2 de dezembro de 2010;31(24):3054–62.

21. Sletten OJ, Aalen JM, Izci H, Duchenne J, Remme EW, Larsen CK, et al. Lateral Wall Dysfunction Signals Onset of Progressive Heart Failure in Left Bundle Branch Block. JACC Cardiovasc Imaging. novembro de 2021;14(11):2059–69.

22. Kuronuma K, Miller RJH, Van Kriekinge SD, Han D, Singh A, Gransar H, et al. Incremental prognostic value of stress phase entropy over standard PET myocardial perfusion imaging variables. Eur J Nucl Med Mol Imaging. 10 de outubro de 2023;50(12):3619–29.

23. Camici PG, Crea F. Coronary Microvascular Dysfunction. New England Journal of Medicine. 22 de fevereiro de 2007;356(8):830–40.

24. Murthy VL, Naya M, Foster CR, Hainer J, Gaber M, Di Carli G, et al. Improved Cardiac Risk Assessment With Noninvasive Measures of Coronary Flow Reserve. Circulation. 15 de novembro de 2011;124(20):2215–24.

25. McGowan RL, Welch TG, Zaret BL, Bryson AL, Martin ND, Flamm MD. Noninvasive myocardial imaging with potassium-43 and rubidium-81 in patients with left bundle branch block. Am J Cardiol. outubro de 1976;38(4):422–8.

26. Botvinick EH, Taradash MR, Shames DM, Parmley WW. Thallium-201 myocardial perfusion scintigraphy for the clinical clarification of normal, abnormal and equivocal electrocardiographic stress tests. Am J Cardiol. janeiro de 1978;41(1):43–51.

27. Nowak B, Sinha AM, Schaefer WM, Koch KC, Kaiser HJ, Hanrath P, et al. Cardiac resynchronization therapyhomogenizes myocardial glucosemetabolism and perfusion in dilatedcardiomyopathy and left bundle branch block. J Am Coll Cardiol. maio de 2003;41(9):1523–8.

28. Salatzki J, Fischer T, Riffel J, André F, Hirschberg K, Ochs A, et al. Presence of contractile impairment appears crucial for structural remodeling in idiopathic left bundle-branch block. Journal of Cardiovascular Magnetic Resonance. março de 2021;23(1):39.

29. Brandão SCS, Nishioka SAD, Giorgi MCP, Chen J, Abe R, Filho MM, et al. Cardiac resynchronization therapy evaluated by myocardial scintigraphy with 99mTc-MIBI: changes in left ventricular uptake, dyssynchrony, and function. Eur J Nucl Med Mol Imaging. 14 de junho de 2009;36(6):986–96.

30. Vernooy K, Cornelussen RNM, Verbeek XAAM, Vanagt WYR, van Hunnik A, Kuiper M, et al. Cardiac resynchronization therapy cures dyssynchronopathy in canine left bundle-branch block hearts. Eur Heart J. 19 de julho de 2007;28(17):2148–55.

31. Hess PL, Shaw LK, Fudim M, Iskandrian AE, Borges-Neto S. The prognostic value of mechanical left ventricular dyssynchrony defined by phase analysis from gated single-photon emission computed tomography myocardial perfusion imaging among patients with coronary heart disease. Journal of Nuclear Cardiology. abril de 2017;24(2):482–90.

