## Supplemental material for "Mechanical Dyssynchrony and Perfusion Heterogeneity Predict Adverse LV Remodeling in Patients with and without LBBB"

### Supplemental Appendix

#### S1. Supplemental Methods

**S1.1 Automated LV Regional Function Assessment:** Regional LV motion (mm; change in endocardial position from end-diastole to end-systole) and wall thickening (%; percent increase in wall thickness in the same interval) were assessed automatically using a validated algorithm (11). The algorithm assigns severity scores for motion (0–5) and thickening (0–3) to five LV regions (septal, lateral, anterior, inferior, apex) based on deviations from a normal population, without requiring visual input. All analyses were performed automatically at rest. Higher scores indicate worsened regional function.

#### S2. Supplemental Results

#### Tables and Figures

##### Table S1. Comparison of regional left ventricular functional indices between control and LBBB groups

| | Groups | | | |
|---|---|---|---|---|
| | **Control (n = 932)** | **LBBB (n = 233)** | | |
| | **Mean ± SD** | **Mean ± SD** | **Mean diff. (95% CI)** | **p Value** |
| **Rest Reg. Mo. (mm)** | Score from 0 to 5; higher scores mean worse regional function. | | | |
| Global | 0.26 ± 0.44 | 0.77 ± 0.77 | -0.52 (-0.62, -0.41) | < 0.001 |
| Anterior | 0.17 ± 0.38 | 0.49 ± 0.65 | -0.32 (-0.41 a -0.23) | < 0.001 |
| Inferior | 0.25 ± 0.50 | 0.72 ± 0.85 | -0.47 (-0.58, -0.35) | < 0.001 |
| Apex* | 0.45 ± 0.66 | 1.28 ± 1.07 | -0.83 (-0.97, -0.68) | < 0.001 |
| Lateral | 0.22 ± 0.40 | 0.48 ± 0.63 | -0.27 (-0.35, -0.18) | < 0.001 |
| Septal* | 0.27 ± 0.65 | 1.24 ± 1.40 | -0.97 (-1.15, -0.78) | < 0.001 |
| **Rest Reg. Thk. (%)** | Score from 0 to 3; higher scores mean worse regional function. | | | |
| Global | 0.05 ± 0.15 | 0.20 ± 0.32 | -0.15 (-0.20, -0.11) | < 0.001 |
| Anterior | 0.03 ± 0.11 | 0.14 ± 0.29 | -0.11(-0.15, -0.07) | < 0.001 |
| Inferior | 0.04 ± 0.13 | 0.16 ± 0.28 | -0.12 (-0.15, -0.08) | < 0.001 |
| Apex* | 0.11 ± 0.30 | 0.50 ± 0.65 | -0.39 (-0.48, -0.31) | < 0.001 |
| Lateral | 0.04 ± 0.11 | 0.11 ± 0.21 | -0.08 (-0.10, -0.05) | < 0.001 |
| Septal* | 0.05 ± 0.18 | 0.26 ± 0.44 | -0.21 (-0.27, -0.15) | < 0.001 |

The mean difference (diff) and corresponding 95% confidence interval (CI) reflect the difference between the control and LBBB groups for each parameter. Negative mean differences mean differences indicate lower values in the control group compared to the LBBB group. LBBB = left bundle branch block; Rest Reg. Mo. = regional endocardial surface motion; Reg. Thk. = regional endocardial wall thickening; SD = standard deviation. *Indicates LV regions with the **most pronounced** worsening of motion and thickening in the LBBB group compared to controls.

**Table S2. Global and regional myocardial blood flow (MBF), myocardial flow reserve (MFR), and coronary vascular resistance (CVR) measurements between control and LBBB groups using PET MPI**

| | **Groups** | | | |
|---|---|---|---|---|
| | **Control** | **LBBB** | | |
| | **Mean ± SD (N =932)** | **Mean ± SD (N =233)** | **Mean diff. (95% CI)** | **p value*** |
| **MBF - mL/min/g** | | | | |
| Rest global MBF | 1.10 ± 0.39 | 1.08 ± 0.37 | 0.02 (-0.04, 0.07) | 0.584 |
| Rest anterior MBF | 1.09 ± 0.40 | 1.09 ± 0.38 | 0.01 (-0.05, 0.06) | 0.794 |
| Rest inferior MBF | 1.08 ± 0.39 | 1.07 ± 0.37 | 0.01 (-0.04, 0.06) | 0.709 |
| Rest apex MBF | 1.10 ± 0.42 | 1.09 ± 0.41 | 0.01 (-0.05, 0.07) | 0.848 |
| Rest lateral MBF | 1.15 ± 0.41 | 1.16 ± 0.41 | -0.01 (-0.07, 0.04) | 0.637 |
| Rest septal | 1.07 ± 0.40 | 0.99 ± 0.36 | 0.08 (0.02, 0.13) | **0.005** |
| Rest SLR | 0.93 ± 0.16 | 0.87 ± 0.24 | 0.06 (0.03, 0.09) | **< 0.001** |
| Stress global MBF | 2.59 ± 0.80 | 2.43 ± 0.77 | 0.16 (0.05, 0.28) | **0.004** |
| Stress anterior MBF | 2.62 ± 0.81 | 2.45 ± 0.75 | 0.17 (0.06, 0.28) | **0.002** |
| Stress inferior MBF | 2.55 ± 0.83 | 2.42 ± 0.81 | 0.13 (0.01, 0.25) | **0.028** |
| Stress apex MBF | 2.44 ± 0.85 | 2.30 ± 0.79 | 0.14 (0.02, 0.25) | **0.023** |
| Stress lateral MBF | 2.66 ± 0.82 | 2.51 ± 0.78 | 0.15 (0.03, 0.26) | **0.012** |
| Stress septal MBF | 2.65 ± 0.86 | 2.40 ± 0.86 | 0.25 (0.12, 0.37) | **< 0.001** |
| Stress SLR | 1.00 ± 0.17 | 0.95 ± 0.18 | 0.05 (0.02, 0.07) | **< 0.001** |

| | | | | |
|---|---|---|---|---|
| **MFR** | | | | |
| Global MFR | 2.55 ± 0.84 | 2.41 ± 0.75 | 0.14 (0.03, 0.25) | **0.013** |
| Anterior MFR | 2.59 ± 0.89 | 2.44 ± 0.85 | 0.15 (0.03, 0.28) | **0.016** |
| Inferior MFR | 2.55 ± 0.91 | 2.42 ± 0.82 | 0.13 (0.00, 0.25) | **0.042** |
| Apex MFR | 2.39 ± 0.87 | 2.24 ± 0.72 | 0.15 (0.04, 0.26) | **0.008** |
| Lateral MFR | 2.51 ± 0.87 | 2.33 ± 0.79 | 0.18 (0.06, 0.29) | **0.003** |
| Septal MFR | 2.67 ± 0.89 | 2.57 ± 0.83 | 0.10 (-0.02, 0.22) | 0.110 |
| S/L MFR ratio | 1.08 ± 0.18 | 1.12 ± 0.23 | -0.04 (-0.07, -0.01) | **0.012** |
| **CVR - mmHg/mL·min$^{-1}$·g$^{-1}$** | | | | |
| Rest global CVR | 92.29 ± 36.84 | 93.57 ± 34.42 | -1.28 (-6.31, 3.75) | 0.616 |
| Rest anterior CVR | 93.35 ± 38.55 | 94.00 ± 35.57 | -0.66 (-5.87, 4.55) | 0.804 |
| Rest inferior CVR | 94.65 ± 39.58 | 95.44 ± 37.23 | -0.79 (-6.22, 4.64) | 0.774 |
| Rest apex CVR | 94.35 ± 41.48 | 94.97 ± 38.38 | -0.62 (-6.24, 5.00) | 0.829 |
| Rest lateral CVR | 88.42 ± 35.44 | 88.31 ± 36.10 | 0.11 (-5.07, 5.29) | 0.968 |
| Rest septal CVR | 97.69 ± 45.43 | 105.14 ± 43.50 | -7.45 (-13.77, -1.13) | **0.021** |
| Rest S/L CVR ratio | 1.10 ± 0.22 | 1.21 ± 0.33 | -0.11 (-0.15, -0.07) | **<0.001** |
| Stress Global CVR | 34.05 ± 13.64 | 36.50 ± 15.12 | -2.45 (-4.60, -0.31) | **0.025** |
| Stress anterior CVR | 33.70 ± 13.52 | 36.05 ± 14.83 | -2.34 (-4.45, -0.23) | **0.030** |
| Stress inferior CVR | 35.08 ± 14.91 | 37.35 ± 17.06 | -2.27 (-4.68, 0.14) | 0.065 |
| Stress apex CVR | 37.41 ± 17.59 | 39.26 ± 17.74 | -1.86 (-4.42, 0.71) | 0.155 |
| Stress lateral CVR | 33.14 ± 13.56 | 34.97 ± 13.79 | -1.83 (-3.82, 0.15) | 0.071 |

| | | | | |
|---|---|---|---|---|
| Stress septal CVR | 34.02 ± 15.50 | 38.80 ± 20.61 | -4.79 (-7.64, -1.94) | **0.001** |
| Stress S/L CVR ratio | 1.03 ± 0.19 | 1.09 ± 0.24 | -0.06 (-0.10, -0.03) | **<0.001** |

LBBB: Left Bundle Branch Block; S/L: Septal-to-Lateral. SLR: Septal-to-Lateral (myocardial blood flow) Ratio. PET MPI: Positron Emission Tomography Myocardial Perfusion Imaging. *Values in bold indicate significance (p<0.05).

**Table S3. Reclassification analysis of the total sample according to both the level of mechanical dyssynchrony (stress phase entropy) and the stress septal-to-lateral myocardial blood flow ratio (SLR)**

| | Total Sample (N = 1161) Subgroups | | | | | |
|---|---|---|---|---|---|---|
| | Stress Phase Entropy < 75th (<52%) n = 870 (75%) | | | Stress Phase Entropy ≥ 75th (≥52%) n = 291 (25%) | | |
| **Subgroups** | **SLR ≥ 1.0** | **SLR < 1.0** | | **SLR ≥ 1.0** | **SLR <1.0** | |
| **n (%)** | 442 (51%) | 427 (49%) | | 105 (36%) | 186 (64%) | |
| **Variables, mean ± SD** | **SLR ≥ 1.0** | **SLR < 1.0** | **p Value** | **SLR ≥ 1.0** | **SLR <1.0** | **p Value** |
| Phase entropy (%) | 36.90 ± 7.94 | 37.69 ± 7.77 | 0.144 | 58.97 ± 5.65 | 60.69 ± 5.98 | 0.01 |
| SLR | 1.13 ± 0.11 | 0.89 ± 0.09 | <0.001 | 1.14 ± 0.15 | 0.84 ± 0.12 | <0.001 |
| LVEF (%) | 69.69 ± 10.82 | 67.60 ± 11.02 | 0.005 | 54.79 ± 16.62 | 49.44 ± 14.93 | 0.007 |
| iEDV (mL/m2 | 39.05 ± 11.88 | 44.40 ± 15.98 | <0.001 | 52.79 ± 23.92 | 60.61 ± 22.40 | 0.007 |
| iESV (mL/m2) | 12.60 ± 7.88 | 15.46 ± 10.36 | <0.001 | 26.69 ± 22.86 | 32.79 ± 19.85 | 0.02 |

The Table shows differences in LVEF, iEDV, and iESV among four groups defined by stress phase entropy (<75th percentile for lower dyssynchrony and ≥75th percentile for higher dyssynchrony) and SLR (<1.0, indicating imbalanced perfusion, and ≥1.0, indicating balanced perfusion).

iEDV: Indexed end-diastolic volume; iESV: Indexed end-systolic volume; LBBB: Left bundle branch block; LVEF: Left ventricular ejection fraction; SD: Standard deviation.

**S2.1 Supplemental Results — Correlation Analyses**

To maintain flow in the main text, detailed association results are reported here. **Supplemental Figure S1** summarizes correlations between mechanical dyssynchrony (stress phase entropy) and perfusion heterogeneity (stress SLR) with LV volumes and function. In LBBB patients, phase entropy showed a positive correlation with iEDV ($r = 0.29$, $p < 0.001$) and iESV ($r = 0.31$, $p < 0.001$), and a negative correlation with LVEF ($r = -0.37$, $p < 0.001$). The stress SLR correlated inversely with iEDV ($r = -0.42$, $p < 0.001$) and iESV ($r = -0.40$, $p < 0.001$), and positively with LVEF ($r = 0.34$, $p < 0.001$). Among controls, significant correlations were also observed between stress phase entropy and iEDV ($r = 0.31$, $p < 0.001$), iESV ($r = 0.43$, $p < 0.001$), and LVEF ($r = -0.52$, $p < 0.001$). Similarly, though with weaker coefficients compared to the LBBB group, the stress SLR was inversely correlated with iEDV ($r = -0.19$, $p < 0.001$) and iESV ($r = -0.15$, $p < 0.001$), and positively with LVEF ($r = 0.12$, $p < 0.001$). Additionally, there was no significant correlation between stress phase entropy and SLR in the control group ($r = -0.04$, $p = 0.163$), whereas a significant inverse correlation was present in the LBBB group ($r = -0.28$, $p < 0.001$).

In both the control and LBBB groups, global stress MBF correlated negatively with iEDV and iESV ($r = –0.34$, $p < 0.001$ for both), but positively with LVEF (control: $r = 0.36$; LBBB: $r = 0.37$; all $p < 0.001$). Conversely, global stress CVR was positively correlated with iEDV and iESV (control: $r = 0.39$ and 0.37; LBBB: $r = 0.36$ and 0.35, for iEDV and iESV, respectively; all $p < 0.001$) and negatively with LVEF (control: $r = –0.36$; LBBB: $r = –0.35$; all $p < 0.001$). MFR showed no significant correlations with these indices in either group ($r = –0.06$ to –0.01 for iEDV/iESV; $r = –0.004$ to 0.008 for LVEF; all $p >0.05$).

**Figure S1. Correlations of stress phase entropy and stress septal-to-lateral myocardial blood flow ratio (SLR) with left ventricular (LV) remodeling and function indices in control and left bundle branch block (LBBB) patients**

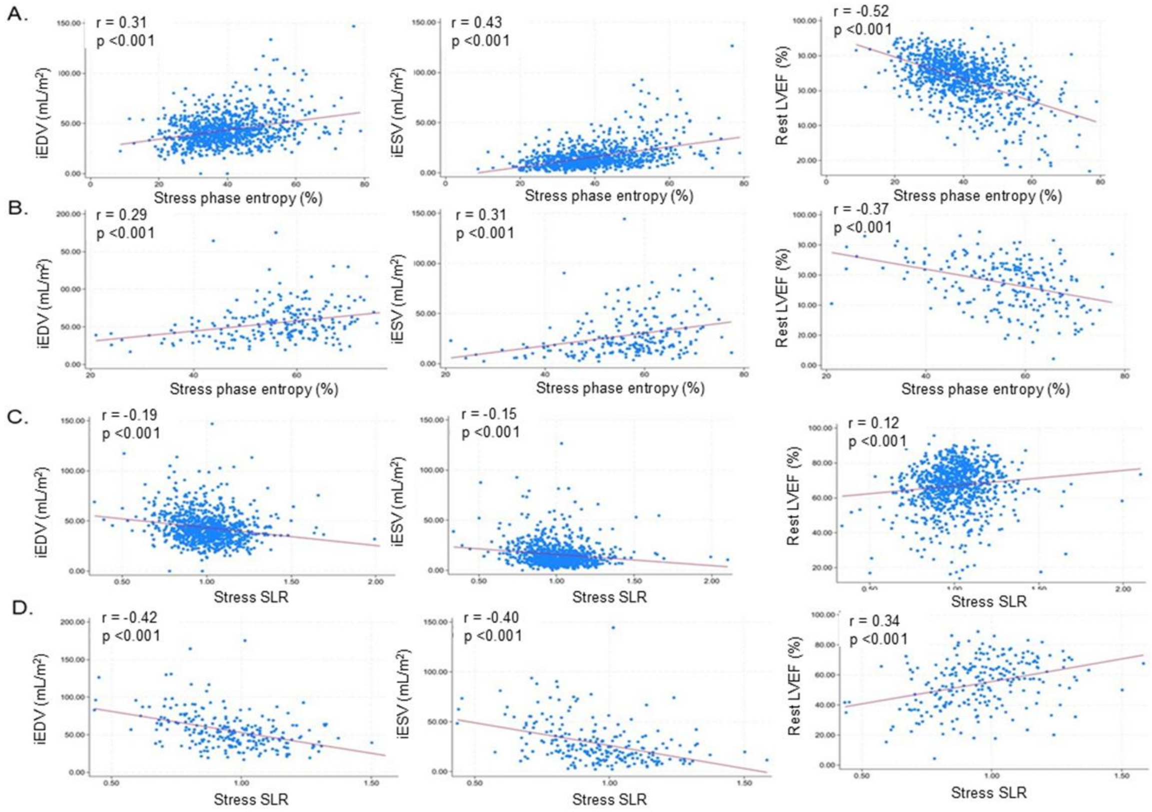


Scatter plots for the control group (A, C) and the LBBB group (B, D). Panels A and B show correlations between **stress phase entropy** and indexed end-diastolic volume (iEDV), indexed end-systolic volume (iESV), and left ventricular ejection fraction (LVEF) at rest. Panels C and D show correlations between **stress SLR** and iEDV, iESV, and LVEF. In both groups, higher stress phase entropy was associated with increased LV volumes and lower LVEF, indicating worse mechanical synchrony and ventricular function. Conversely, a higher stress SLR was associated with smaller LV volumes and higher LVEF, reflecting less adverse remodeling and better ventricular function.
